# Co-designing an intervention to strengthen vaccine uptake in Congolese migrants in the UK (LISOLO MALAMU): a participatory study protocol

**DOI:** 10.1101/2022.07.19.22277798

**Authors:** Alison F Crawshaw, Caroline Hickey, Laura Muzinga Lutumba, Lusau Mimi Kitoko, Sarah Luti Nkembi, Felicity Knights, Yusuf Ciftci, Lucy P Goldsmith, Tushna Vandrevala, Alice S Forster, Sally Hargreaves

**Author notes:** **Corresponding author** Sally Hargreaves, The Migrant Health Research Group, Institute for Infection and Immunity, St George’s, University of London, Cranmer Terrace, London, SW17 0RE, UK.

## Abstract

**Introduction:** Adult migrants are at risk of under-immunisation and are likely to need catch-up vaccination to bring them in line with the UK schedule. The COVID-19 pandemic has highlighted and exacerbated inequities in vaccine uptake, with migrants facing additional barriers to information, low vaccine confidence, and access to vaccine services. There is a need for participatory and theory-based research that meaningfully engages underserved migrant groups to make sense of their experiences and beliefs about vaccination and uses these insights to co-produce tailored interventions which can increase uptake. COVID-19 vaccination provides a unique entry-point and opportunity to explore these issues in tandem with addressing routine immunisation gaps and developing more culturally-sensitive routine vaccination services.

**Methods and analysis:** LISOLO MALAMU (‘Good Talk’) is a community-based participatory research study which uses co-design, design thinking and behaviour change theory to engage adult Congolese migrants in developing a tailored intervention to increase vaccine uptake. A community-academic coalition will lead and co-design the study. The study will involve i) in-depth interviews with adult Congolese migrants (foreign-born, >18 years), ii-iii) interviews and consensus workshops with clinical, public health and community stakeholders, and iv) co-design workshops with adult Congolese migrants. Qualitative data will be analysed iteratively, using Thematic Analysis, and mapped to the Theoretical Domains Framework, with participation from the coalition in discussing and interpreting findings and selecting intervention functions to guide the co-design workshops. Sociodemographic data of interview participants will be summarised using descriptive statistics. The study will run from approximately November 2021-November 2022.

**Ethics and dissemination:** Ethics approval has been granted by the St George’s University Research Ethics Committee (REC reference 2021.0128). Study findings will be widely disseminated by the coalition through local community organisations in Hackney and broader academic and policy stakeholders, including a final celebration event. Recommendations for a future larger scale study and testing of prototyped interventions will be made.

**Strengths and limitations of this study:** 

**Strengths:**

- This study will directly respond to ongoing calls for community-centred and participatory approaches to engaging migrants in routine and COVID-19 vaccination, by implementing a value-driven and reciprocal approach to conducting a study addressing the needs of an underserved community.
- The target population was selected following a comprehensive systematic review of the evidence (1) and pre-engagement scoping work conducted with migrant community representatives in London, UK. (2, 3)
- It aims to co-produce a tailored intervention to address specific barriers to, and strengthen, vaccine uptake for COVID-19 and routine vaccines in adult Congolese migrants (including MMR, Td/IPV, and HPV) as set out by UKHSA guidance (4), and has been co-designed with, and will be co-delivered by, a coalition formed of academic researchers, a council for voluntary service (a local charity which offers services and support for local voluntary and community organisations), and a Congolese community-based organisation.

**Limitations:**

- As this study is tailored to the Congolese migrant population, other migrants who also face barriers to vaccine uptake are not included. Whilst we can draw some conclusions about the experiences of other Black migrants who face similar historical and cultural barriers to uptake of routine and COVID-19 vaccines, our ability to generalise the findings to all migrant communities might be limited.
- Co-designed intervention prototypes will not be formally implemented and evaluated in this study, however recommendations will be made so that this can be done in a future phase.

## Introduction

Adult migrants (foreign-born individuals) in Europe, particularly those from low-and middle-income countries, are at risk of under-immunisation for routine vaccinations (5-8) and have been involved in outbreaks of serious vaccine-preventable diseases, including measles (9). The reasons for their undervaccination are multiple and complex, and include differing vaccination schedules in migrants’ home countries, practical factors relating to access and availability, historical and cultural reasons, and other individual and social processes, which can occur before, during, and after migration (1, 10, 11).

Unlike children, who are typically aligned with the host country’s vaccination schedule upon attending school, adult and adolescent migrants are not routinely incorporated into vaccination programmes on arrival to most European countries, including the UK (12), due to lack of guidance, and well-documented barriers to accessing health systems D. Our recent systematic review (1) confirmed that access barriers including language, literacy, communication, practical, legal, and service barriers, are particularly important barriers to vaccination for migrants in transit and host countries, and that specific factors including country of origin (particularly African), having more recently migrated, and being an asylum seeker or refugee, could be determinants of under-immunisation in migrants.

The World Health Organization has launched its new Immunization Agenda 2030 (IA2030) (13) with an emphasis on achieving equitable access to vaccination for vulnerable populations and integrating vaccination throughout the lifecourse, including catching-up adolescent and adult migrants with missed vaccines, doses and boosters, to close immunisation gaps. WHO and ECDC guidance for catch-up vaccination is available (14, 15) and in the UK specific guidance on the ‘vaccination of individuals with uncertain or incomplete immunisation status’ is available from UKHSA (16), although the implementation of this guidance in practice is unknown and thought to be inconsistent, with low awareness of the guidance by clinicians in primary care (17).

Besides routine vaccination, there have been striking disparities in COVID-19 vaccine uptake, with UK and US evidence showing migrants and minoritised populations – particularly Black groups – having some of the lowest uptake rates to date. (18-20) The COVID-19 pandemic has exposed inequities in engaging adult migrants and other under-served populations in vaccination programmes (11, 21) and highlighted how structural racism and marginalisation serve to perpetuate their poorer health outcomes (22). If not adequately addressed, the enduringly low levels of COVID-19 vaccine uptake in these populations are likely to widen existing inequities. A lack of adult immunisation programmes worldwide – and therefore crucial infrastructure for delivering routine and COVID-19 vaccinations globally – also poses a barrier to achieving equitable COVID-19 vaccine deployment and uptake among adults, and must be addressed (23).

Emerging evidence, including from our pre-engagement work done to inform this study (reported elsewhere (2, 3)) attributes lower COVID-19 vaccine uptake rates in minoritised and migrant populations to a lack of confidence – specifically due to the spread of misinformation and conspiracy theories, mistrust in the medical establishment and government, and concerns about side effects – and access barriers, including physical access, language and communication barriers (9, 24-29). In the UK, community-research partnerships have been mobilised to engage with ethnic communities and address the disparities in COVID-19 vaccine uptake including through outreach and the development of culturally relevant health information and messages, (30, 31) and public bodies and charities have now translated official information and guidance into multiple languages and developed toolkits (32-34). However, there are fewer initiatives to use community partners as equal partners in research and community-based participatory research (CBPR) studies specifically engaging and involving migrant populations around vaccination, and indeed on other health topics. (35) To date, few studies have closely and carefully explored the barriers and facilitators in specific migrant sub-populations with inequitable uptake, and for whom mainstream interventions and resources have failed to reach or influence, with the purpose of using these insights to co-produce tailored interventions. This is amid calls from the World Health Organization (WHO) and the European Centre for Disease Prevention and Control (ECDC) for more participatory research on the determinants of health in migrant populations, and better understanding of the needs and perspectives of refugees and migrants in vaccination initiatives (36, 37). There is a real opportunity for the renewed focus on engaging underserved populations brought about by COVID-19 to be carried over to routine vaccinations, with improved inclusion of migrant populations, who continue to be overlooked.

CBPR approaches, which aim to equitably involve all partners in the research process (38), hold potential to tackle this complex issue because they recognise the value of lived experience and emphasise the sharing of power, ensuring research is embedded within, conducted in collaboration with, and tailored to, a specific community (39, 40). Behaviour change models and frameworks, such as the Theoretical Domains Framework, can be used in tandem to understand the context of behaviour and design targeted interventions (41), and have been applied in a variety of health settings, including vaccination (42). The relevance of CBPR approaches to migrant health research has been noted (43) and evidence shows that interventions driven by insights from the communities they are designed to serve are more cost-effective and lead to better results for health behaviour outcomes than traditional interventions (44, 45). A systematic review which looked at strategies for addressing vaccine hesitancy globally found that multi-component and dialogue-based interventions were most effective, and recommended that strategies should be carefully tailored to the target population, their specific reasons for hesitancy, and context (46). Recent research around COVID-19 testing with people of Black ethnicity in the UK highlighted the role of mistrust, alienation and stigmatisation in creating barriers to testing for this population, and recommended that health communications address these issues and build trust through local credible sources (47). Evidence also supports the importance of locality in establishing and maintaining trust and credibility in the processes of community engagement and health promotion, and for developing contextually-specific, tailored interventions (47).

To inform this study, we conducted a systematic review of the literature on migrant vaccine uptake in Europe (1) (including local grey literature) and 3 online pre-engagement workshops with migrant community representatives in London (in December 2020-February 2021), to scope out community perceptions towards COVID-19 vaccination and barriers and facilitators to uptake, and found support within communities to participate in research to co-develop solutions (2, 3). Based on our key findings that recent migrants, refugees and asylum seekers, and those from Africa may be at risk of being under-immunised (1), local data showing that people from the Black community in Hackney have some of the lowest rates of vaccine coverage for routine childhood vaccination and COVID-19 vaccination (48, 49), evidence of widespread access and confidence barriers affecting migrants’ COVID-19 vaccine uptake locally, and the relationships built with community organisations during our pre-engagement work, a community-academic partnership was formed and adult Congolese migrants (predominantly from the Democratic Republic of Congo, DRC) were chosen as our target population. The DRC migrant population in the UK has a large proportion of older adult migrants who began migrating to the UK in the late 1980s, many as political refugees, with increased flows since the late 1990s (50). DRC refugees were the fourth most common nationality to be resettled to the UK through the UK’s four main refugee resettlement schemes between 2015-2020 (51) (n=1774) and recent unpublished data suggest that UK-bound adult DRC refugees may be significantly under-immunised compared to the UK immunisation schedule (52). The Congolese population in the UK is historically underserved and there is very limited literature on their health-seeking behaviour and health outcomes. This study will therefore make an important contribution to the evidence base.

The aim of this study is to use CBPR approaches to engage and involve Congolese migrants in Hackney in the co-design of a tailored intervention to increase vaccine uptake. It seeks to i) gather information about and make sense of Congolese adult migrants’ beliefs, experiences of vaccination (routine, COVID-19, catch-up), access to healthcare, and other lived experiences with respect to vaccination, ii) understand local pathways, processes and services, and considerations for implementation of interventions with key stakeholders; and iii) co-design a tailored intervention to strengthen vaccine uptake with Congolese migrants, which can be formally evaluated.

## Methods and analysis

### Study design

LISOLO MALAMU (Lingala for ‘Good Talk’) is a CBPR study which uses co-design methods, the principles of design thinking (an iterative, solutions-based approach to problem-solving that starts with the needs and desires of the target population) (53) and behaviour change theory (41) to engage Congolese migrants in developing a tailored intervention to increase vaccine uptake. The comprehensive and practical Theoretical Domains Framework (TDF) and accompanying Behaviour Change Wheel (BCW), specifically developed for implementation research, were chosen to guide the intervention design in this study (41). The study name was decided in consultation with the community to reflect the project’s ethos of providing a platform for meaningful conversations around vaccination. It involves 4 main activities: 1) community days, involving qualitative in-depth interviews with Congolese migrants, which began in January 2022, 2) in-depth interviews (IDIs) with local clinical, public health and community stakeholders, 3) consensus workshops with the same key stakeholders, and 4) co-design workshops with Congolese migrants. An evaluation component will be embedded across all activities. The study process is illustrated in Figure 1. Good practices, challenges and facilitators relating to the implementation of the study and the method of using co-design will also be documented. An academic-community research partnership has been formed (referred to herewith as “the coalition”) to co-design, steer and conduct the study.

**Figure 1.** Study process and activities, mapped to the 4 design thinking phases of empathise (with target population); define (target population’s needs, their problems and your insights); ideate (by challenging assumptions and creating ideas for innovative solutions); prototype (to start creating solutions). (53)

### Setting and population

The study is being carried out in Hackney, London, UK, a highly diverse London borough, in which more than 89 languages are spoken and around 40% of the population come from Black and Minority Ethnic Groups. (54) It was the 11^th^ most deprived local authority in England in the Indices of Deprivation 2015. (55) The study will be conducted with adult migrants (>18 years) predominantly from the DRC and with local clinical, public health and community stakeholders based in Hackney. Specific inclusion and exclusion criteria are described in Table 1. There are an estimated 23,000 migrants from the DRC and Republic of Congo combined living in the UK, with Hackney hosting one of the largest communities. (56)

**Table 1.** Inclusion and exclusion criteria.

| Target population | Inclusion criteria | Exclusion criteria |
| --- | --- | --- |
| Migrants | Born in the Democratic Republic of the Congo (Congo-Kinshasa), Republic of Congo (Congo-Brazzaville), Angola, or another Lingala-speaking region of Central Africa.<br>Aged 18 or above.<br>Currently residing in the UK.<br>Willing and able to give informed consent. | Not migrant as per earlier definition.<br>Not born in one of the specified countries/regions.<br>Below the age of 18.<br>Temporarily in the UK for holiday, visiting friends/family or other reasons.<br>Individuals who may lack the capacity to consent, as determined by the mental capacity act framework. |
| Local stakeholders | Aged 18 or above.<br>Volunteer or employee of a local group, organisation or business that has a vested interest in the health of the target community, such as local government, public health, National Health Service (NHS), community, and faith-based organisations.<br>Willing and able to give informed consent. | Not a local stakeholder as per earlier definition.<br>Below the age of 18.<br>Individuals who may lack the capacity to consent, as determined by the mental capacity act framework. |

### Study team and coalition

A coalition was formed in November 2021 to steer the study including 3 members of Hackney Congolese Women Support Group (HCWSG) (LML, LMK, SN - 2 Congolese migrant women and 1 British woman of Congolese descent), 1 network coordinator from Hackney Council for Voluntary Service (HCVS) (CH, a British woman with a Master’s in Community Engagement and extensive community and voluntary sector experience), 1 lead researcher from St George’s, University of London (AFC, a White migrant woman with an MSc in Control Infectious Diseases and extensive experience in social and behaviour change communication and implementation research), and 3 other academic co-researchers who provide an advisory function. The researchers will facilitate the involvement of the coalition members, providing research training and helping them to understand and contribute to the research process. HCWSG will facilitate engagement with the local Congolese community and HCVS will facilitate relationships with the local integrated care system (a collaborative partnership between the organisations that deliver health and care needs locally) and voluntary and community sector.

### Support for partners

Study partners from HCWSG and HCVS were financially compensated for their time and effort (57). All study resources and expenses were paid for by the project budget managed by the St George’s research group. Non-financial contributions to HCWSG included honorary membership to the School of Oriental and African Studies (SOAS) Library, London (as requested to access African and Congolese literature to support their community work), training and upskilling opportunities, and grantwriting support.

### Planning

In November-January 2021, the coalition had three 2-hour meetings to assign roles and responsibilities, plan the study, map the target population and stakeholders, and refine the research questions and approach, one half-day training session on qualitative interviewing techniques, led by AFC, and one half-day session to practice, pilot test and refine the interview topic guide. Based on the community’s preference for oral communication and face-to-face interactions, the coalition decided that the study should be promoted mostly by word of mouth and flyers co-designed by the coalition, and that data from the Congolese community should be collected face-to-face (COVID-19 restrictions permitting).

### Recruitment

The study seeks to recruit approximately 30 migrants living in and around Hackney, London, UK, to participate in the semi-structured qualitative interviews, 6-8 migrants to participate in the co-design workshops, and approximately 4-6 local (to Hackney) stakeholders to participate in the key informant interviews and consensus workshops, although actual sample sizes will be guided by data saturation. Participants will be recruited through publicity among the coalition’s networks (e.g., by email bulletins, word of mouth, community meetings and advertisements) and by additional snowball sampling techniques. Participants will be compensated according to NIHR guidance (57) and reasonable expenses (travel, childcare, etc) will be paid.

### Data collection

The study data and data collection methods are described in Table 2. Interviews with migrants will be conducted by 4 members of the coalition in Lingala, French or English, depending on the participant’s preference (LML, LMK, SN are trilingual; AFC speaks English and will use an interpreter as required). Interviews with local stakeholders will be conducted in English by the lead researcher; consensus workshops will be co-facilitated by the coalition in English and co-design workshops will be co-facilitated by the coalition in Lingala, French and English. Qualitative interview data will be collected iteratively with a pilot-tested topic guide, which will be revised and refined as data are generated. Discussions with the interviewers will take place regularly to review the interview guide and data collection.

**Table 2.** Study activities, data, and data collection methods.

| No. | Activity | Population | Data generated/collected | Data collection methods |
| --- | --- | --- | --- | --- |
| 1 | Community days (n~3) | Congolese migrants (n~30) living in and around Hackney, London, UK | Beliefs and experiences related to vaccination<br>Suggestions for engagement approaches and interventions<br>Community values<br>Sociodemographic information | IDIs (n~30)<br>Post-it notes/interactive posters/graffiti walls<br>Sociodemographic surveys |
| 2 | Key informant interviews | Local clinical, public health and community stakeholders (n~6) | Role and relationship with the Congolese community<br>Description of local pathways, processes, and services<br>Suggestions for potential interventions and considerations for implementation | IDIs |
| 3 | Consensus workshops (n~2) | Local clinical, public health and community stakeholders (n~6) | Insight and evidence-based discussion to generate feedback and suggestions for intervention design | Consensus workshops |
| 4 | Co-design workshops (n~2) | Congolese migrants (n~8) | Feedback and iteration on intervention prototypes | Participatory workshops |
| n/a | Evaluation | All populations plus community coalition | Feedback on involvement in co-design process<br>Feedback on participation in study activities (IDIs, workshops) | Evaluation forms/questionnaires<br>Voting |
|  |  |  | Feedback on final prototype |  |

#### Activity 1

Approximately 30 semi-structured, in-depth qualitative interviews with Congolese migrants will be conducted by the coalition to explore beliefs, perceptions and experiences relating to catch-up vaccination for routine vaccinations including MMR, Td/IPV, HPV, flu, and COVID-19 vaccination, and obtain suggestion for novel interventions. These will be delivered through “community days” held at HCVS, which is closely located to the local market that the Congolese community attend for their weekly shopping. Community days were planned to coincide with market days to facilitate attendance. To date, two community days have been held at HCVS, with interviews conducted in private rooms and a central social area provided for the community to gather over Congolese food and music. Additional data and insight about Congolese culture and values were collected through interactive posters in the social space. Post-interview evaluation forms and sociodemographic surveys were also collected. Information about other local community services (e.g. educational classes) were provided and referrals were facilitated by the HCVS coalition member.

#### Activity 2

Approximately 4-6 in-depth, online interviews will be conducted with local key informants/stakeholders (e.g. local GPs/nurses, clinical and public health staff, religious leaders and relevant community organisations in Hackney), to explore their role and relationship with the Congolese community, understand local pathways, processes and services and discuss potential interventions and considerations for implementation.

#### Activity 3

1-2 online consensus workshops will be conducted with local stakeholders (from activity 2) to discuss emerging findings and obtain feedback and suggestions to inform ongoing data collection and design of interventions.

#### Activity 4

Approximately 2 co-design workshops will be conducted in-person with two groups of 4-6 Congolese migrants who participated in the in-depth interviews (activity 1) to discuss and iterate on the intervention functions that were selected by the coalition following data synthesis and appraisal and create an intervention prototype.

#### Evaluation

Activities will be evaluated with feedback from participants and community feedback on the final intervention prototype will be sought at the celebration event.

### Analysis and preparation of initial intervention prototypes

Qualitative interview and consensus workshop data will be analysed iteratively, using Thematic Analysis (58), in NVivo software (Mac version). Anonymised digital recordings will be translated into English and transcribed verbatim by an independent professional translator, and transcripts, field notes, anonymous evaluation forms and other data collected during the activities (post-it notes, posters) will be imported into NVivo for coding and analysis. Sociodemographic data will be entered into Excel, aggregated, and summarised using descriptive statistics. Qualitative and quantitative analysis will be led by SGUL researchers, in consultation with the coalition to discuss, member-check, triangulate and interpret findings and define emergent themes. Themes will be mapped to the TDF (59) and BCW (60) to identify behavioural components and potential intervention functions (defined as broad categories of means by which an intervention can change behaviour) needed to change behaviour (41). Following data synthesis, interpretation and analysis from activities 1-3, the lead researcher will prepare a short summary of key findings and the corresponding intervention functions identified from the BCW to present to and verify with the coalition. The coalition will consider the candidate intervention functions using the APEASE criteria (affordability, practicability, effectiveness/cost effectiveness, acceptability, side-effects/safety, equity) (41), discuss potential interventions employing these functions that could be effective and tailored to the target population, and decide by consensus on approximately 2 intervention functions to take forward to the co-design workshops with Congolese community members. These intervention functions will be the starting point for the workshops, and potential intervention strategies involving these functions will be discussed, iterated on, and tailored with the participation of the community, with the end goal being to co-produce a single, detailed intervention prototype. Any summary notes from the workshops and photographs of visual data generated (e.g. post-it notes, illustrations, etc) will subsequently be imported into NVivo software for data management and further analysis by the coalition.

### Schedule

The planned duration of the study is 12 months, starting from November 2021 and ending in November 2022.

### Patient and public involvement

The initial idea for this study was informed by informal scoping workshops with diverse migrant community representatives and community-based organisations in London (predominantly City & Hackney) that were conducted by St George’s, University of London in collaboration with Hackney CVS in January-March 2021 and published (2, 3). These participants will be invited to a roundtable discussion of findings and a dissemination event at the end of the study. The coalition was established to co-design and deliver the study. The study name was chosen by the coalition in consultation with the community to reflect the project’s ethos of providing a platform for meaningful conversations around vaccination in the Congolese community. The planned consensus workshops with local stakeholders (activity 3) and co-design workshops with Congolese migrants (activity 4) will directly involve members of the public in designing tailored intervention prototypes. An independent patient and public involvement board (St George’s Migrant Health Research Group NIHR Project Board) comprising 5 adult migrants with lived experience of accessing healthcare in the UK will also be consulted at significant points over the course of the study.

## Ethics and dissemination

This study has been given favourable ethical opinion by the St George’s University Research Ethics Committee (REC reference 2021.0128). A celebration event and webinar for participants, the local community and key stakeholders will be organised at the end of the study. The study findings will be widely disseminated at local, national and international levels, including conferences, policy and stakeholder meetings, voluntary and community sector assemblies, peer-reviewed journals, a PhD thesis, and multimedia outputs, e.g. video clips and tweets. Research data and outputs will be stored in the St George’s Research Data Repository. Recommendations for a future larger scale study and testing of prototyped interventions will be made.

## Supporting information

SRQR Reporting Checklist

## Data Availability

The manuscript is a prospective study protocol and therefore does not contain data, however the data collected in future by the study will be available upon reasonable request to the authors.

## Acknowledgments

We thank the Hackney Congolese Women Support Group, Hackney Refugee and Migrant Forum and Hackney CVS, members of St George’s Migrant Health Research Group NIHR Project Board, and the community representatives and community organisations consulted during our pre-engagement work. We thank all the participants in our study and in particular the Congolese communities in Northeast London.

## Authors’ contributions

AFC wrote the first draft, with input from SH and ASF, and all authors reviewed and commented on a final version.

